# Automatic approach-avoidance tendency toward physical activity, sedentary, and neutral stimuli as a function of age, explicit affective attitude, and intention to be active

**DOI:** 10.1101/2022.09.05.22279509

**Authors:** Ata Farajzadeh, Miriam Goubran, Alexa Beehler, Noura Cherkaoui, Paula Morrison, Margaux de Chanaleilles, Silvio Maltagliati, Boris Cheval, Matthew W. Miller, Lisa Sheehy, Martin Bilodeau, Dan Orsholits, Matthieu P. Boisgontier

**Affiliations:** School of Rehabilitation Sciences, Faculty of Health Sciences, University of Ottawa, Canada; Bruyère Research Institute, Ottawa, Canada; Laboratory Sport and Social Environment (SENS), Université Grenoble Alpes, France; Swiss Center for Affective Sciences, University of Geneva, Switzerland; Laboratory for the Study of Emotion Elicitation and Expression (E3Lab), Department of Psychology, University of Geneva, Switzerland; School of Kinesiology, Auburn University, USA; Center for Neuroscience, Auburn University, USA; Centre for the Interdisciplinary Study of Gerontology and Vulnerability, University of Geneva, Switzerland; Swiss National Centre of Competence in Research LIVES—Overcoming Vulnerability: Life Course Perspectives, Lausanne and Geneva, Switzerland

**Keywords:** Aging, Attitude, Exercise, Geriatrics, Health, Intention, Personality, Reaction Time, Sedentary Behavior

## Abstract

Using computerized reaction-time tasks assessing automatic attitudes, studies have shown that healthy young adults have faster reaction times when approaching physical activity stimuli than when avoiding them. The opposite has been observed for sedentary stimuli. However, it is unclear whether these results hold across the lifespan and when error rates and a possible generic approach-avoidance tendency are accounted for. Here, reaction times and errors in online approach-avoidance tasks of 130 participants aged 21 to 77 years were analyzed using mixed-effects models. Automatic approach-avoidance tendencies were tested using physical activity, sedentary, and neutral stimuli. Explicit attitudes toward physical activity and intention to be physically active were self-reported. Results accounting for age, sex, gender, level of physical activity, body mass index, and chronic health condition confirmed a main tendency to approach physical activity stimuli (i.e., faster reaction to approach vs. avoid; p = .001) and to avoid sedentary stimuli (i.e., faster reaction to avoid vs. approach; p < .001). Results based on neutral stimuli revealed a generic approach tendency in early adulthood (i.e., faster approach before age 53 and fewer errors before age 36) and a generic avoidance tendency in older adults (i.e., more errors after age 60). When accounting for these generic tendencies, results showed a greater tendency (i.e., fewer errors) to avoid than approach sedentary stimuli after aged 50, but not before (p = .026). Exploratory analyses showed that, irrespective of age, participants were faster at approaching physical activity (p = .028) and avoiding sedentary stimuli (p = .041) when they considered physical activity as pleasant and enjoyable (explicit attitude). However, results showed no evidence of an association between approach-avoidance tendencies and the intention to be physically active. Taken together, these results suggest that both age and explicit attitudes can affect the general tendency to approach physical activity stimuli and to avoid sedentary stimuli.

---

Since the pioneering work on mental chronometry (Donders, 1868) and the first time the term “Reactionszeit” was used (Exner, 1873), reaction time has been a means to study brain function. In particular, reaction-time tasks can reveal what psychologists call implicit or automatic attitudes, defined as “introspectively unidentified traces of past experience that mediate favorable or unfavorable feeling, thought, or action toward a social object” (Greenwald & Banaji, 1995). In other words, an automatic attitude is thought to result from the positive or negative value that our brain automatically assigns to some concept (e.g., person, place, or behavior), without that value being accurately accessible to cognition (however, see Corneille & Hutter, 2020 for a critical discussion). This implicit value of a stimulus results in an automatic positive or negative inclination toward this stimulus, which influences behavior.

## Automatic Attitudes & Health Behavior

Automatic attitudes to health-related stimuli are thought to influence health behavior (Marteau et al., 2012). Beyond correlational evidence, some intervention studies targeting these attitudes, called cognitive bias modification interventions, have proven they can be successful in changing health behavior (Kakoschke et al., 2017). For example, interventions have been used to retrain the automatic reaction to alcohol (Wiers et al., 2011; Eberl et al., 2013; Gladwin et al., 2015; Rinck et al., 2018). Using a joystick, participants were repeatedly asked to avoid pictures on a screen that were related to alcohol and to approach pictures related to soft drinks. Results showed this intervention reduced the alcohol relapse rate by 9% to 13% one year after treatment discharge (Wiers et al., 2011; Eberl et al., 2013; Gladwin et al., 2015). This type of intervention has also proven to be useful in altering smoking (Wittekind et al., 2015) and eating behavior (Aulbach et al., 2019; Kemps et al., 2019) as well as anxiety and depressive disorders (Taylor & Amir, 2012; Fodor et al., 2020). However, the effectiveness of these intervention has also been questioned (Becker et al., 2018; Brockmeyer et al., 2019).

In physical activity, automatic attitudes have been shown to be associated with behavior (Chevance et al., 2019), but causality has not yet been demonstrated in ecological settings. A recent pilot intervention study including 40 students (20 per arm) showed no evidence of an effect of an intervention aimed at increasing physical activity through the modification of automatic attitudes (Preis et al., 2021). However, the absence of significance is not evidence of the absence of effect (Harms & Lakens, 2018), especially since a power calculation from a recent protocol article estimated that a minimum of 252 participants (126 per arm) would be needed to demonstrate efficacy of this type of intervention with a probability of committing a type I error <5% and a probability of committing a type II error <10% (Cheval et al., 2021b). Further, a laboratory study showed that a single session of automatic attitude training could influence physical activity behavior (Cheval et al., 2016b). Taken together, these studies suggest that automatic attitudes are associated with health behaviors, including physical activity, and provide some evidence for a causal relationship.

### Reaction Time & Conceptual Congruence

To study automatic attitude, researchers analyze the reaction time to the simultaneous (e.g., implicit association test [Greenwald et al., 1998]) or sequential (e.g., affective priming [Fazio et al., 1986]) presentation of a reference stimulus (e.g., a neutral stimulus) and an experimental stimulus of interest (e.g., an image of physical activity). If the participant’s brain evaluates, based on the accumulation of information from past experiences, that the concepts carried by the reference and experimental stimulus are congruent, the time required to process and react to the experimental stimulus will be shorter than if it were presented with a neutral or incongruent reference stimulus. Using this approach, researchers can determine whether the stimulus of interest is congruent with a positive reference stimulus (e.g., a positive word) suggesting a positive automatic attitude toward the stimulus of interest or, conversely, whether it is congruent with a negative reference stimulus (e.g., a negative word), suggesting a negative automatic attitude. From a sensorimotor perspective, the extensive psychology literature using this approach (Greenwald et al., 2009) unambiguously demonstrates that reaction time depends not only on the number and complexity of stimuli to be processed (Donders, 1869), but also on the conceptual congruence between the concepts carried by the stimuli or between a stimulus and the concept carried by an action toward that stimulus (e.g., to approach vs. to avoid).

### Automatic Approach-Avoidance Tendency

Conceptual congruence can also be revealed by manipulating the physical (e.g., pulling or pushing a joystick) or virtual direction (e.g., pressing keyboard keys moving an avatar on a screen; selecting the word “approach”) of the response used to react to the stimulus of interest (Krieglmeyer & Deutsch, 2010; Rougier et al., 2018). While generic approach-avoidance tendencies have been studied using questionnaires at the personality (e.g., approach-avoidance temperaments) (Elliott et al., 2002) and goal level (approach-avoidance goals) (Carver & White, 1994; Elliott, 1999), the reaction-time difference in these approach-avoidance tasks captures a more automatic aspect of approach-avoidance tendencies, a specific dimension of automatic attitude (Sheeran et al., 2013). In a seminal study, Solarz showed that reaction times were faster when participants pulled cards with pleasant words toward themselves and when they pushed cards with unpleasant words away from themselves rather than the reverse (Solarz, 1960). Since then, this effect suggesting an automatic tendency to approach positively-valued concepts and avoid negatively-valued concepts has been replicated numerous times with various types of approach-avoidance tasks and across numerous contexts (Chen & Bargh, 1999; Wentura et al., 2000; De Houwer et al., 2001; Duckworth et al., 2002; Vaes et al., 2003; Rotteveel & Phaf, 2004; Markman & Brendl, 2005; Alexopoulos & Ric, 2007; Rinck & Becker, 2007; Paladino & Castelli, 2008; Seibt et al., 2008; Saraiva et al., 2013; Rougier et al., 2018). As approach-avoidance tendencies play a key role in adapting a broad range of behaviors to the perception of one’s context (Lang, 1995), this construct has attracted considerable attention in physical activity.

### Aging & Physical Activity

In exercise and sports science, studies based on these approach-avoidance tasks have consistently shown faster reaction times when approaching physical activity stimuli and avoiding sedentary stimuli (Cheval et al., 2014; Cheval et al., 2015; Cheval et al., 2016a; Cheval et al., 2018c; Hannan et al., 2019; Moffit et al., 2019; Locke & Berry, 2021). These results suggest a positive evaluation of the concept of physical activity and a negative evaluation of the concept of sedentary behavior. However, these studies were conducted in healthy, young to middle-aged adults, most often kinesiology students. To the best of our knowledge, whether age affects this tendency to approach physical activity and avoid sedentary behavior has not been tested. Yet, the age-related increase in perceived physical fatigability (LaSorda et al., 2020) and chronic pain (Shupler et al., 2019) may contribute to an increase in the number of situations where physical activities are associated with unpleasant experiences across aging. This accumulation of negative experiences related to physical activity could potentially reduce automatic positive attitudes toward this behavior. Concurrently, sedentary behaviors may become more attractive (Maher & Dunton, 2020; O’Brien et al., 2021). Therefore, results from healthy, younger populations may not be generalizable to older populations. This is an important knowledge gap to fill since the world’s population of people aged 60 years and older will double between 2015 and 2050 to 2.1 billion (World Health Organization, 2021). A better understanding of the determinants of the age-related decline in physical activity will contribute to reduce the risk of disability (Martin Ginis et al., 2021), chronic diseases (Bauer et al., 2014), and mortality (Saint-Maurice et al., 2021) as well as the economic burden of over $67 billion per year (Ding et al., 2016) associated with physical inactivity. Low physical activity levels among older adults, reaching 60% of the population in the Americas (Hallal et al., 2012), as well as their decline across aging (Cheval et al., 2018b; Cheval et al., 2020b), make the study of automatic attitudes toward physical activity and sedentary behaviors even more important in this population. Such research would complement the growing literature that examines deliberative factors involved in regulating these behaviors (e.g., explicit attitudes, intention) (Maartje et al., 2009; Koeneman et al., 2011).

### Explicit Attitudes & Intentions

Automatic attitudes were originally conceived as independent of explicit self-reported measures such as explicit attitudes and intentions (Norman & Shallice, 1986). Yet, recent findings suggest that they can influence each other and partially overlap (Greene et al., 2001; Hofmann et al., 2005; Nosek, 2005; Béna et al., 2022). In kinesiology, studies based on the implicit association test investigating the association of automatic attitudes toward physical activity with explicit attitudes and intention to be physically active have shown inconsistent results. Some studies showed a positive association of automatic attitudes with explicit attitudes (Muschalik et al., 2019) and intentions (Banting et al., 2009), whereas other studies found no evidence of these associations (Conroy et al., 2010; Hyde et al., 2010; Rebar et al., 2015; Sala et al., 2016; Muschalik et al., 2018). Similarly, a study testing automatic approach-avoidance tendencies showed an association between automatic attitudes and intention (Cheval et al., 2015), whereas other studies did not (Cheval et al., 2014; Hannan et al., 2021). However, none of these studies examined error rates, which could have confounded the results due to the tendency for decision speed to covary with accuracy (speed-accuracy trade-off) (Hick, 1952; Heitz, 2014). Because faster reactions are more error-prone than slower reactions, individuals may be faster with lower accuracy or slower with higher accuracy (Chittka et al., 2009). Accordingly, interpreting reaction times without considering errors can be misleading because a reaction time with a high error rate and the same reaction time with a low error rate cannot be considered the same behavior. In addition, most of these studies did not properly assess automatic attitudes toward sedentary behavior, as sedentary stimuli were often considered as a baseline condition against which reaction times to physical activity stimuli were compared. Yet, sedentary behavior is independently related to health (Stamatakis et al., 2019) and its determinants remain poorly understood (Chastin et al., 2015; Maltagliati et al., 2022). Finally, these previous studies did not account for a potential generic approach-avoidance tendency, independent of the type of stimuli used, that could vary across participants, potentially confounding the results. For example, when an individual approaches physical activity stimuli faster than they avoid them, one might conclude there is an automatic tendency to approach these stimuli. However, this individual may also have a generic tendency to approach rather than avoid any stimuli. Therefore, not controlling for this generic tendency could lead to the erroneous conclusion that an individual has an automatic tendency to approach physical activity stimuli, when in fact they automatically tend to approach any stimulus. Accounting for potential inter-individual differences in generic approach-avoidance tendencies is even more important when including participants of different ages, as personality research has shown that younger adults reported higher approach motivation compared to midlife and older adults (Windsor et al., 2012).

### Hypothesis & Objectives

As preregistered (Boisgontier, 2021), the hypotheses tested in this study were that the automatic tendency to avoid sedentary behavior increases with age (i.e., faster reaction times or fewer errors to approach versus avoid sedentary stimuli) and that the automatic tendency to engage in physical activity behavior decreases with age (i.e., slower reaction times or more errors to approach versus avoid physical activity stimuli). In addition, exploratory analyses were conducted to test whether automatic approach-avoidance tendencies toward physical activity and sedentary behaviors were associated with explicit attitudes and the intention to be physically active across aging.

## Methods

### Population

Participants were recruited through social media (Facebook), posters at the Faculty of Health Sciences, University of Ottawa, and word-of-mouth. Inclusion criteria were age 20–80 years and access to a personal computer or a laptop with internet. Participants who did not complete the full study or used a phone or tablet were excluded. Informed consent was collected in accordance with the Declaration of Helsinki. The study was approved by University of Ottawa’s Research Ethics Boards (H-05-21-6791). All participants provided informed consent. Data were collected between December 2021 and June 2022. Participants were not compensated for their participation.

An a priori power analysis was conducted in G*power (Faul et al., 2009) to estimate the minimum sample required for α = 0.05, power (1-β) = 90%, and a medium effect size f^2^ = 0.15 (Cohen, 1988). The analysis was based on an F test in the linear multiple regression (R^2^ increase) that included the highest number of predictors (six tested predictors including two interaction effects and a total of eleven predictors) estimated that a minimum sample size of n=123 was required. We expected that 14% of the participants would fail the attention check questions (Steele et al., 2021). Therefore, we planned to recruit at least 144 participants.

### Experimental Protocol

#### Procedures

Participants performed approach-avoidance tasks online using Inquisit 6 software (Millisecond Software, 2015), and responded to questions related to their age, sex (male, female), gender (man, woman, non-binary, transgender man, transgender woman, other), weight, height, explicit attitude toward physical activity, intention to be physically active, usual level of physical activity, and chronic health condition. Two attention check questions were included in the questionnaires based on this model: “Please answer “2” to this question that allows us to verify that you actually read the questions.”

#### Approach-Avoidance Task

Automatic approach-avoidance attitudes toward physical activity and sedentary stimuli were tested using an approach-avoidance task because it allowed the intensity of both positive and negative automatic attitudes to be derived from the assessment of approach-related and avoidance-related behaviors, respectively, rather than only the semantic aspects of attitudes (Znanewitz et al., 2018). Further, this task has shown good reliability (splithalf method: r = 0.76) (Rinck & Becker, 2007), which was similar to the reliability of an approach-avoidance implicit association test (ρ = .77) (Moffit et al., 2020). In terms of validity, the approach-avoid task has shown the most consistent pattern of associations with outcomes related to physical activity (Zenko & Ekkekakis, 2019b).

Automatic tendencies to approach or avoid physical activity behavior and sedentary behavior were assessed using the approach-avoidance task in two experimental conditions and two neutral conditions (Cheval et al., 2018c). In the experimental conditions of this task, a trial starts with a fixation of a cross presented at the center of the screen for a random time ranging from 500 to 750 ms (Figure 1A). Then, an avatar appears either at the top or bottom third of the screen for one second, before a pictogram representing a physical activity behavior or a sedentary behavior appears in the center of the screen (Figure 1A). The participant sitting in front of the computer with one index finger positioned on the “U” and the other index finger on the “N” key is instructed that pressing the “U” key moves the avatar up and pressing the “N” key moves the avatar down. Accordingly, the movement of the avatar is always congruent with the pressed key: The top key (i.e., U) moves the avatar up, while the bottom key (i.e., N), moves the avatar down. Importantly, however, the approach or avoidance action depends on the initial position of the avatar at the beginning of the trial. If the avatar appears below the stimulus, the top key is associated with an approach movement, while the bottom key is associated with an avoidance movement. Conversely, if the avatar appears above the stimulus, the approach and avoidance movement are reversed – the top key is associated with an avoidance movement and the bottom key is associated with an approach movement. This design provides the manikin task an advantage over the joystick tasks, as explained in the seminal work by Krieglmeyer and Deutsch: “Contrary to the joystick tasks, in the manikin task (De Houwer et al., 2001) recategorization is rather unlikely. Although in principle, the movements can be recategorized as moving downwards and upwards, this would make the task more difficult instead of easier. The reason for this is that the manikin either appears above or below the stimulus, and, therefore, up and down responses are unrelated to the instructed approach-avoidance responses. Thus, depending on the position of the manikin, participants have to determine in each trial which response means approach or avoidance. Consequently, the representation of approach and avoidance is activated in each trial.”

**Figure 1.**
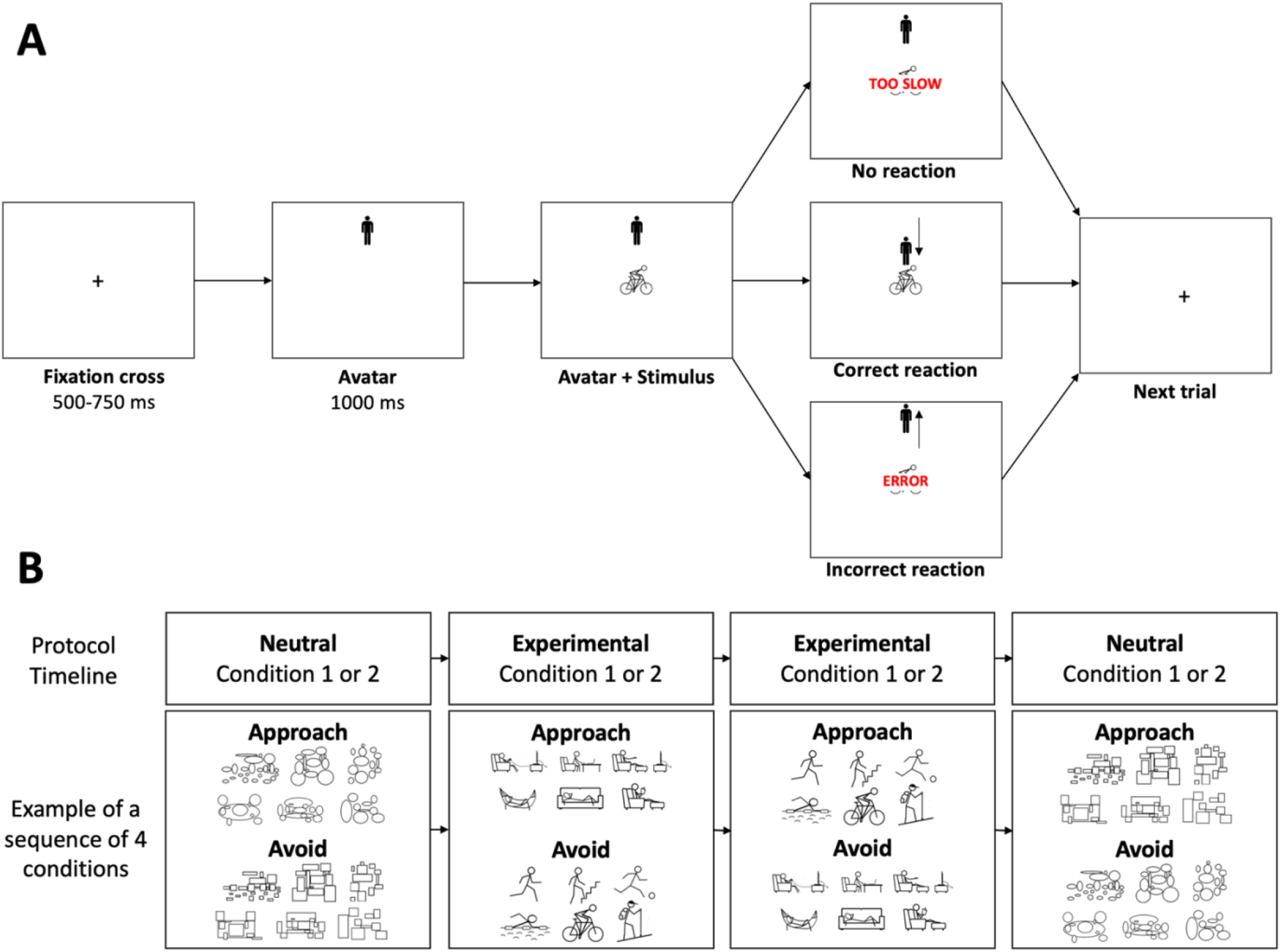
A. Illustration of a trial of the approach-avoidance task in the condition where the participant is instructed to approach physical activity stimuli (and avoid sedentary stimuli – not shown). B. Timeline and stimuli of the approach-avoidance task. In the experimental and neutral Condition 1, the participant is instructed to move the avatar toward (i.e., approach) a type of stimuli (i.e., physical activity or rectangles) and to move the avatar away from (i.e., avoid) stimuli depicting the other type of stimuli (i.e., sedentary behavior or ellipses, respectively). In Condition 2, the instruction is reversed: The participant is instructed to move away from physical activity (experimental condition) or rectangle stimuli (neutral condition) and move toward sedentary stimuli or ellipse stimuli.

Two experimental conditions were tested (Figure 1B). In one experimental condition, the participant is instructed to quickly move the avatar toward (i.e., approach) pictograms depicting physical activity and to move the avatar away from (i.e., avoid) pictograms depicting sedentary behavior. In the other experimental condition, the participant does the opposite: move away from physical activity and move toward sedentary stimuli. The order of the experimental conditions was randomized across participants.

In addition to the two experimental conditions, two neutral conditions were tested. These conditions were used to account for a potential generic approach-avoidance tendency that could vary across participants and ages (Windsor et al., 2012). In these neutral conditions, the stimuli depicting physical activity and sedentary behaviors were replaced by stimuli made of rectangles or ellipses that matched the number and size of information in 3 physical activity stimuli (swimming, hiking, cycling) and 3 sedentary stimuli (couch, hammock, reading). Two conditions were tested. In one condition, participants are asked to quickly move the manikin toward stimuli with circles and away from stimuli with squares. In the other condition, the participant is given opposite instructions. The order of the neutral conditions was randomized.

One neutral condition was tested before the two experimental conditions, and the other neutral condition was tested after them. Each condition included 96 stimuli, 48 of each class (physical activity and sedentary stimuli in the experimental conditions; rectangles and ellipses in the neutral conditions), that were presented randomly. Familiarization with the task was performed during the first 15 trials of the study, which were removed from the analyses. Familiarization with the subsequent conditions was performed during the first 3 trials of each condition, which were removed from the analyses. The physical activity and sedentary stimuli were presented all together on the screen for seven seconds before each experimental condition. Between conditions, the participant could rest for as long as they wanted before pressing the space key to start the next condition. When the participant pressed the incorrect key (“U” when it should be “N” or “N” when it should be “U”), the message “error” appeared on the screen for 800 ms before the next trial. When the reaction time (i.e., the time between the appearance of the stimuli and the key press) was longer than seven seconds, the message “too slow” appeared on the screen for 800 ms before the next trial (Figure 1A).

The automatic tendency to approach or avoid a type of stimuli (i.e., physical activity, sedentary, or neutral stimuli) was derived from the time required to press the key in reaction to a type of stimulus (i.e., physical activity vs. sedentary vs. neutral). Incorrect responses, responses faster than 150 ms, and responses slower than 3,000 ms were excluded from the analyses to account for outliers and loss of attention. This latter threshold is double what is recommended in young adults (Krieglmeyer & Deutsch, 2010) because a 1,500-ms threshold would have resulted in a loss of 20.8 % of observations (vs. 3.4 % with the 3,000 ms threshold), primarily in the older participants, which could have biased the results.

As recommended (Zenko and Ekkekakis, 2019a), we estimated the internal consistency of bias using a permutation-based splithalf approach (splithalf package; Parsons et al., 2019; Parsons, 2022) with 5,000 random splits. The Spearman-Brown corrected splithalf internal consistency (r_SB_) of reaction-time bias toward physical activity, sedentary, neutral, and all stimuli was 0.68 (95% confidence intervals [95CI] = 0.55 – 0.78), 0.70 (95CI = 0.59 – 0.79), 0.88 (95CI = 0.83 – 0.92), and 0.83 (95CI = 0.77 – 0.88), respectively. The Spearman-Brown corrected splithalf internal consistency (r_SB_) of error bias toward physical activity, sedentary, neutral, and all stimuli was 0.77 (95CI = 0.68 – 0.84), 0.62 (95CI = 0.46 – 0.74), 0.77 (95CI = 0.68 – 0.84), and 0.75 (95CI = 0.64 – 0.82), respectively. Some authors have suggested that r_SB_ be interpreted as follows: < 0.50 = “poor” reliability, [0.50 – 0.75 [= “moderate” reliability, [0.75 – 0.90] = “good” reliability, and > 0.90 = “excellent” reliability (Koo & Li, 2016). Accordingly, internal consistency of the approach-avoidance task was good when all conditions were included and was moderate to good when each of the three condition was considered separately.

#### Physical Activity and Sedentary Stimuli

In a previous study (Cheval et al., 2018), thirty-two participants were asked to rate the extent to which 24 stimuli expressed “movement and an active lifestyle” and “rest and sedentary lifestyle” (1 = not at all, 7 = a lot). For each stimulus, the “rest and sedentary lifestyle” score was subtracted from the “movement and active lifestyle” score. In the current study, the six stimuli with the largest positive and negative differences were chosen as the stimuli depicting physical activity and sedentary behaviors, respectively.

#### Intention to Be Physically Active

The intention to be physically active was derived from the response to the question “How much do you agree with the following statements: Over the next 7 days, I intend to do at least 150 minutes of moderate-intensity physical activity; or at least 75 minutes of vigorous intensity physical activity; or an equivalent combination of moderate- and vigorous-intensity physical activity” on a 7-point scale ranging from “strongly disagree” (1) to “strongly agree” (7). Due to the skewed distribution of the scores (Supplemental Figure 1), this variable was dichotomized in responses below 7 (N = 76) and responses equal to 7 (N = 54).

#### Explicit Affective Attitude Toward Physical Activity

Explicit attitudes toward physical activity were calculated as the mean of two items (Cronbach’s alpha = 0.92) based on two bipolar semantic differential adjectives on a 7-point scale (unpleasant-pleasant; unenjoyable-enjoyable). The statement begins with “For me, to participate in regular physical activity is …” (Hoyt et al., 2009). Due to the skewed distribution of the scores (Supplemental Figure 2), this variable was dichotomized in responses below 7 (N = 79) and responses equal to 7 (N = 51).

#### Usual Level of Moderate-to-Vigorous Physical Activity

The usual level of physical activity was derived from the short form of the International Physical Activity Questionnaire (IPAQ-SF). The IPAQ-SF is a self-administered questionnaire that identifies the frequency and duration of moderate and vigorous physical activity, as well as sedentary time during the past 7 days to estimate usual practice of physical activity and sedentary behavior (Craig et al., 2003). The usual level of moderate-to-vigorous physical activity (MVPA) in minutes per week was used as a control variable in the analyses.

#### Chronic Health Condition

The absence or presence of a chronic health condition was derived from the question “Has a doctor ever told you that you had any of the following conditions?” based on item PH006 of the Survey of Health, Ageing and Retirement in Europe (Börsch-Supan, 2022). The response “None” was coded as no chronic health condition. The other responses were coded as presence of chronic health condition.

### Statistical Analyses

Data were analyzed using linear and logistic mixed-effect models. This statistical approach is often preferred to traditional analyses such as ANOVAs (Boisgontier & Cheval, 2016) because it avoids information loss due to averaging over participants and increases the number of data points in the model (Judd et al., 2012), which reduces type 1 error rate without compromising statistical power (Baayen et al., 2008). In addition, it allows incomplete and unbalanced data to be used, as well as continuous and categorical predictors to be combined. Here, the mixed-effects models were built and fit by maximum likelihood in the R software environment (R Core Team, 2021), using the lme4 (Bates et al., 2021) and lmerTest package (Kuznetsova et al., 2020), which approximates *p*-values using Satterthwaite’s degrees of freedom method. Continuous variables were standardized. For linear mixed-effects models, restricted maximum likelihood (REML) was used as it provides less biased estimates of variance components than full maximum likelihood (Luke, 2017). When some observations were suspected to exert undue influence on the model estimation, the models were tested with and without them to ensure robustness of the results. An estimate of the variance explained by a fixed effect of interest was reported by subtracting the marginal pseudo R^2^ (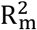 computed with the MuMIn package [Barton, 2022]) of the model without the effect from the 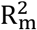 of the model including this effect. 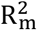 is dimensionless and independentof sample size (Nakagawa et al., 2017), which makes it ideal to compare effect sizes of different models. For the computation of 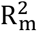, maximum likelihood was used to make 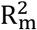 comparable across models with different fixed effects.

#### Statistical Analysis of Reaction Times

Seven linear mixed-effects models used reaction time as outcome. To investigate the effect of age on approach-avoidance bias toward the different stimuli, the first three models tested the interaction effect between age (continuous) and action direction (approach vs. avoid) on reaction time to physical activity stimuli (Model 1), sedentary stimuli (Model 2), and neutral stimuli (Model 3). Models 1, 2, and 3 were not merged in a single model including stimulus (physical activity, sedentary behavior, neutral) as a factor because we were interested in whether age moderated reaction time to approach vs. avoid a specific stimulus, not whether age moderated the effect of stimulus on reaction time to approach versus avoid. In addition, we did not have sufficient statistical power to add a triple interaction to the tested models (age × action direction × stimulus interaction), which would have tested 19 predictors in the same model. The models were tested with different age centrations to determine the age range during which the effects of interest were significant.

As the results of Model 3 showed an effect of age on approach-avoidance bias toward neutral stimuli, a generic approach-avoidance bias could have confounded the results. To account for this potential confounder, corrected reaction times were computed by subtracting the mean reaction time to approach or avoid neutral stimuli from the reaction time on each trial to approach or avoid stimuli depicting a type of behavior (sedentary or physical activity behavior), respectively. Then, a model was conducted to test the interaction effect between age and action direction on corrected reaction time to physical activity (Model 4; Equation 1) and sedentary stimuli (Model 5; Equation 1).

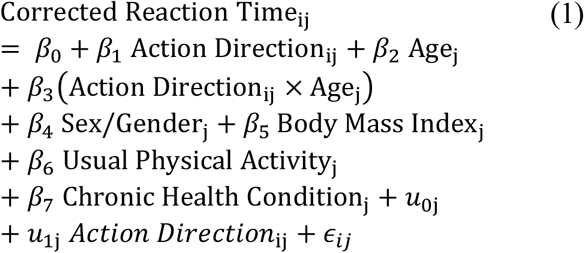

In this equation, *Corrected Reaction Time*_*ij*_ is the j^th^ participant’s corrected reaction time on trial *i*, the *β*s are the fixed effect coefficients, *u*0j is the random intercept for the *j*^th^ participant, *u*1j is the random slope of the action direction condition for the *j*^th^ participant, and *ϵ*_*ij*_ is the error term.

The last two models tested if the explicit attitude toward physical activity and the intention to be physically active showed an interaction effect with action direction on corrected reaction time to physical activity (Model 6; Equation 2) and sedentary stimuli (Model 7; Equation 2) independent of age. All models specified participants and action direction as random factors and were adjusted for sex-gender, body mass index, usual physical activity, and chronic health condition. However, to ensure the robustness of the results, all the models were also conducted without adjusting for these variables. Moreover, the effects of explicit attitude and intention were also tested in separate models.

#### Statistical Analysis of Errors

To ensure that the results obtained with reaction times cannot be explained by the speed-accuracy trade-off (Hick, 1952; Heitz, 2014), nine logistic mixed-effects models used error as outcome. The structure of these models was similar to the linear mixed-effects models using reaction time as outcome (2.4.1). Specifically, the structure of Model 1, 2, 3, 4, and 5 (outcome = reaction time) was used to build Model 1.2, 2.2, 3.2, 4.2, and 5.2 (outcome = error), respectively. However, because the logistic mixed-effects models did not converge when sex-gender, body mass index, usual physical activity, and chronic health condition were included, these variables were removed. In addition, because the models including both explicit attitude and intention did not converge, these variables were tested in separate models. Therefore, the structure of Model 6 was used to build Model 6.2 (explicit attitude) and 6.3 (intention). The structure of Model 7 was used to build Model 7.2 (explicit attitude) and 7.3 (intention). Finally, due to the binary nature of the outcome (error vs. no error), we could not use the same procedure as for reaction time to account for a possible generic approach and avoidance tendency. Instead, the models were adjusted for the mean error of each participant in the condition of avoidance or approach of neutral stimuli.

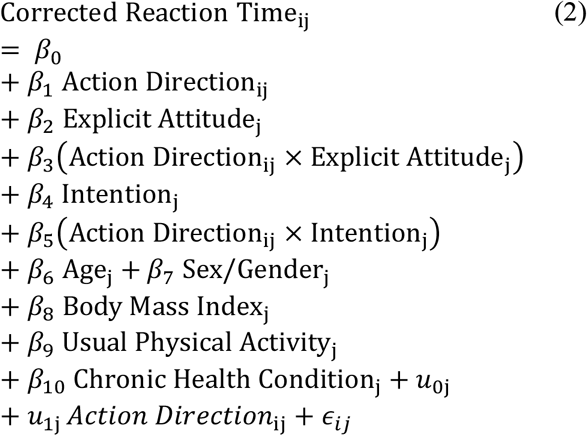

## Results

### Descriptive Results

#### Sample Participants

Two hundred thirty-eight volunteers initiated the study. One hundred sixty-nine participants completed the study (69 stopped the session before completing the study, either due to lack of motivation or technical problems). Some participants were excluded because they answered the check questions incorrectly (n = 3), used a phone or a tablet (n = 30), did not report (n = 3) or reported aberrant height or weight (n = 3) resulting in a final sample of 130 participants (mean age ± SD = 48.2 ± 16.9 years; age range = 21-77 years) with 20 to 24 participants per 10-year age ranges (Supplemental Figure 3). Mean body mass index of the sample was 25.7 ± 4.8 kg/m^2^, the mean usual level of moderate-to-vigorous physical activity was 383.1 ± 445.0 min per week, and 70 participants reported having a chronic health condition. All male (n = 58) and female participants (n = 72) identified themselves as men and women, respectively.

#### Observations

A total of 22,089 reactions times were collected from the participants who reported their age, sex-gender, height, weight, usual physical activity, and chronic health condition. Among these observations, 3.4 % were > 3,000 ms (n = 765), 0.2 % were < 150 ms (n = 58), and 6.7 % were incorrect responses (n = 1,484). Supplemental Table 1 details these observations by condition. A total of 19,971 observations were included in the linear mixed-effects models that have reaction time as outcome (5,358 observations for physical activity stimuli; 5,311 observations for sedentary stimuli; 9,302 observations for neutral stimuli). A total of 21,266 observations was included in the logistic mixed-effects models that had error or no error as outcome (5,654 observations for physical activity stimuli; 5,644 observations for sedentary stimuli; 9,968 observations for neutral stimuli).

#### Descriptive Statistics in Younger, Middle-Aged, and Older Adults

Table 1 presents descriptive statistics of the mean reaction times to approach or avoid the stimuli in young, middle-aged, and older adults. The mean time to approach physical activity stimuli and to avoid sedentary stimuli was faster than the mean time to avoid and approach these stimuli, respectively, in all three age ranges. The mean time to approach neutral stimuli was faster in young and middle-aged adults but was slower in older adults. Explicit affective attitude toward physical activity decreased with age (5.8 ± 1.2 in young adults, 5.6 ± 1.4 in middle-aged adults, and 5.5 ± 1.8 in older adults). The intention to be physically active decreased with age (5.5 ± 1.6 in young adults, 4.9 ± 2.1 in middle-aged adults, and 4.8 ± 2.3 in older adults). The usual level of moderate-to-vigorous physical activity was the lowest in middle-aged adults (223 ± 196 min per week) and the highest in older adults (623 ± 567 min per week), with young adults in between (326 ± 415 min per week). Body mass index were ≥ 25 kg/m^2^ (25.0 ± 5.2 kg/m^2^ in young adults, 26.5 ± 4.2 kg/m^2^ in middle-aged adults, and 25.6 ± 5.0 kg/m^2^ in older adults). Supplemental Table 2 presents correlations of the reaction time to approach or avoid the stimuli (physical activity, sedentary, and neutral stimuli) with age, explicit affective attitude, intention to be active, usual level of moderate-to-vigorous physical activity, and body mass index.

**Table 1.** Descriptive statistics in younger, middle-aged, and older adults. Mean reaction times are in ms, usual physical activity in min per week, and body mass index in kg/m^2^.

| | 21-39 years of age<br>($n = 46$; 23 women; 17 with a chronic health condition) | | | 40-59 years of age<br>($n = 44$; 23 women; 22 with a chronic health condition) | | | 60-77 years of age<br>($n = 40$; 26 women; 31 with a chronic health condition) | | |
| --- | --- | --- | --- | --- | --- | --- | --- | --- | --- |
|  | Mean | SD | Range | Mean | SD | Range | Mean | SD | Range |
| Physical activity stimuli – Approach | 733 | 323 | 321 – 2967 | 937 | 436 | 344 – 2973 | 1281 | 569 | 216 – 2993 |
| Physical activity stimuli – Avoid | 856 | 384 | 339 – 2859 | 1017 | 411 | 214 – 2909 | 1316 | 538 | 219 – 2979 |
| Sedentary stimuli – Approach | 817 | 383 | 304 – 2875 | 1043 | 484 | 266 – 2980 | 1442 | 620 | 378 – 2990 |
| Sedentary stimuli – Avoid | 813 | 353 | 214 – 2985 | 1009 | 418 | 293 – 2962 | 1272 | 504 | 324 – 2927 |
| Neutral stimuli – Approach | 685 | 341 | 247 – 2745 | 851 | 402 | 156 – 2996 | 1231 | 601 | 217 – 2999 |
| Neutral stimuli – Avoid | 736 | 356 | 235 – 2996 | 920 | 426 | 215 – 2996 | 1186 | 541 | 184 – 2983 |
| Explicit affective attitude | 5.8 | 1.2 | 3 – 7 | 5.6 | 1.4 | 1 – 7 | 5.5 | 1.8 | 1 – 7 |
| Intention to be active | 5.5 | 1.6 | 2 – 7 | 4.9 | 2.1 | 1 – 7 | 4.8 | 2.3 | 1 – 7 |
| Usual level of physical activity | 326 | 415 | 0 – 2520 | 223 | 196 | 0 – 780 | 623 | 567 | 0 – 2160 |
| Body mass index | 25.0 | 5.2 | 16.2 – 45.9 | 26.5 | 4.2 | 18.4 – 36.3 | 25.6 | 5.0 | 18.5 – 46.0 |

### Age

#### Age & Physical Activity Stimuli: Uncorrected Results

Model 1 (outcome = reaction time) showed a significant interaction effect between age and action direction (approach vs. avoid) on reaction time to physical activity stimuli (b = 39.9; 95CI = 6.7 – 73.1; 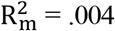; *p* = .020) (Supplemental Table 3). A simple effect analysis revealed that the effect of age is more pronounced in the approach condition than in the avoid condition (b = 230.3; 95CI = 179.7 – 280.9; *p* = 8.4 × 10^−15^ vs. b = 190.4; 95CI = 141.2 – 239.5; *p* = 1.1 × 10^−11^) (Figure 2A). Participants were significantly faster at approaching than avoiding physical activity stimuli until 64 years of age.

**Figure 2.**
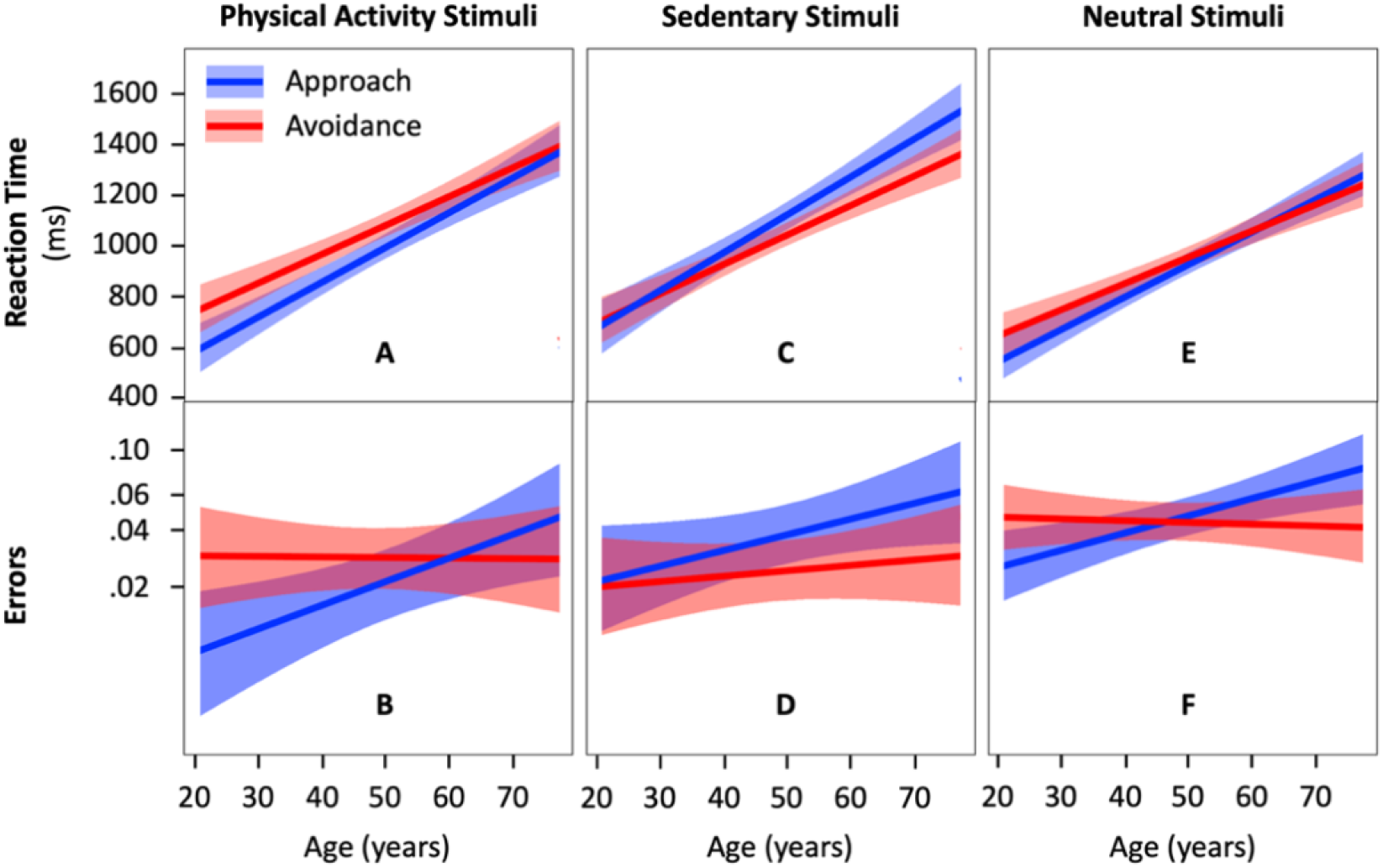
Reaction time to approach and avoid physical activity (A), sedentary (C), and neutral stimuli (E) across adulthood (n = 130 participants) and the corresponding errors (B, D, and F, respectively). The colored area around the regression lines represents the 95% confidence interval.

Model 1.2 (outcome = error) showed a significant interaction effect between age and action direction (approach vs. avoid) on error in physical activity condition (b = 0.500; 95CI = 0.090 – 0.911; 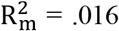 ; *p* = .016) (Supplemental Table 4). A simple effect analysis revealed that the effect of age is more pronounced in the approach condition than in the avoid condition (b = 0.493; 95CI = 0.137 – 0.848; *p* = .006 vs. b = -0.007; 95CI = -0.318 – 0.304; *p* = .963) (Figure 2B). Participants made fewer errors when approaching than avoiding physical activity stimuli until 41 years of age.

Results of Model 1 and Model 1.2 are consistent as they show faster reaction times and fewer errors when approaching compared to avoiding physical activity stimuli before 45 years of age.

#### Age & Sedentary Stimuli: Uncorrected Results

Model 2 (outcome = reaction time) showed a significant interaction effect between age and action direction on reaction time in the condition with sedentary stimuli (b = 56.9; 95CI = 21.4 – 92.6; 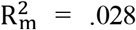*p* = .002) (Supplemental Table 3). A simple effect analysis revealed that the effect of age is more pronounced in the approach condition than in the avoid condition (b = 252.0; 95CI = 196.1 – 308.2; *p* = 1.4 × 10^−14^ vs. b = 195.1; 95CI = 147.5 – 242.7; *p* = 1.2 × 10^−12^) (Figure 2C). Participants were significantly faster at avoiding than approaching sedentary stimuli from age 40 onwards.

Model 2.2 (outcome = error) showed no evidence of an interaction effect between age and action direction on error in the sedentary condition (b = 0.201; 95CI = -0.127 – 0.530; 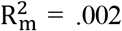:*P* = .229) (Figure 2D; Supplemental Table 4). Similarly, results showed no evidence of a main effect of action direction (b = 0.422; 95CI = -0.025 – 0.870; 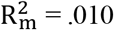 ; *p* = .064).

#### Age & Neutral Stimuli: A Generic Approach-Avoidance Bias

Model 3 (outcome = reaction time) showed a significant interaction effect between age and action direction on reaction time to neutral stimuli (b = 40.7; 95CI = 23.1 – 58.3; 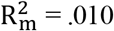; *p* = 1.5 × 10^−5^) (Supplemental Table 5). A simple effect analysis revealed that the effect of age is more pronounced in the approach condition than in the avoid condition (b = 215.8; 95CI = 172.3 – 259.3; *p* < 2.0 × 10^−16^ vs. b = 175.0; 95CI = 131.1 – 218.9; *p* = 7.8 × 10^−4^) (Figure 2E). Participants were significantly faster at approaching than avoiding neutral stimuli until 52 years of age.

Model 3.2 (outcome = error) showed a significant interaction effect between age and action direction on error in the neutral condition (b = 0.391; 95CI = 0.200 – 0.582; 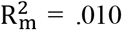; *p* = 6.0 × 10^−5^) (Supplemental Table 6). A simple effect analysis revealed that the effect of age is more pronounced in the approach condition than in the avoid condition (b = 0.358; 95CI = 0.139 – 0.578; *p* = .001 vs. b = -0.032; 95CI = -0.241 – 0.183; *p* = .767) (Figure 2F).Participants made fewer errors when approaching than avoiding neutral stimuli until 35 years of age. From age 36 to 57, errors to approach and avoid neutral activity stimuli were not statistically different. From age 58 onward, participants made statistically more errors when approaching than avoiding neutral stimuli.

#### Age & Physical Activity Stimuli: Results Accounting for a Generic Approach-Avoidance Bias

Because the interaction between age and action direction observed in Model 1, 1.2, and 2 can be explained by a generic effect of age on the tendency to approach stimuli as suggested by the results of Model 3 and Model 3.2, the remaining models (Model 4 to 7 and 4.2 to 7.3) account for this potential confounder (see section 2.4).

Model 4 (outcome = reaction time) showed no evidence of an interaction between age and action direction on corrected reaction time to physical activity stimuli (b = 0.1; 95CI = -36.6 – 37.0; 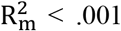 ; ;*p* = .992) (Table 2). However, results showed a significant main effect of action direction (b = -60.3; 95CI = -97.1 – -23.3; 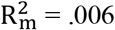 ; ;*p* = .001), with faster reactions to approach than avoid physical activity stimuli (Figure 3A).

**Table 2.**
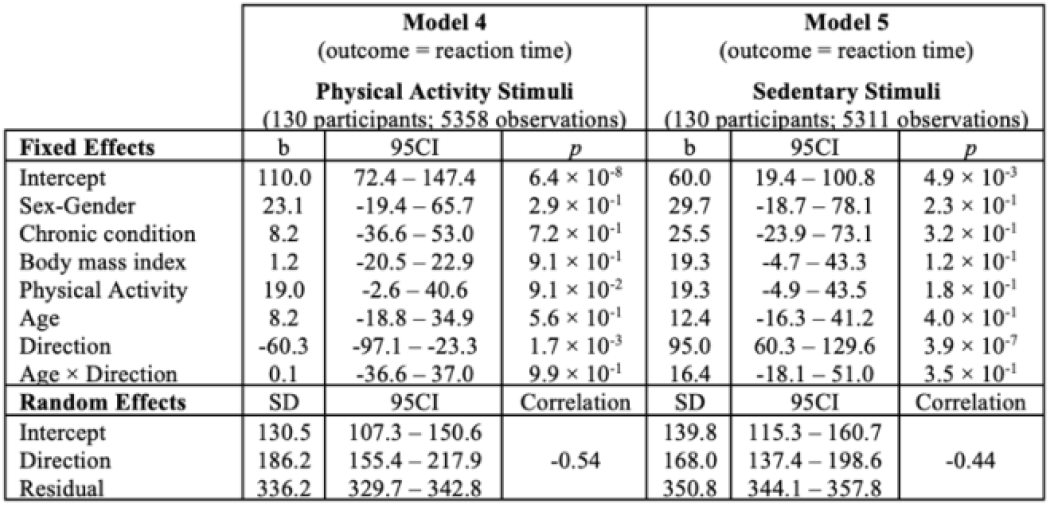
Estimated interaction effect of age and action direction on the reaction time to physical activity (Model 4) and sedentary stimuli (Model 5) corrected for the mean reaction time to neutral stimuli. 95CI = 95% confidence interval.

Model 4.2 (outcome = error) showed no evidence of an interaction between age and action direction on error in the physical activity condition (b = 0.197; 95CI = -0.214 – 0.608; 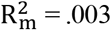 ; *p* = .347) and no evidence of a significant main effect of action direction (b = -0.321; 95CI = -0.803 – 0.160; 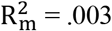 ; *p* = .191) (Table 3; Figure 3B).

**Table 3.**
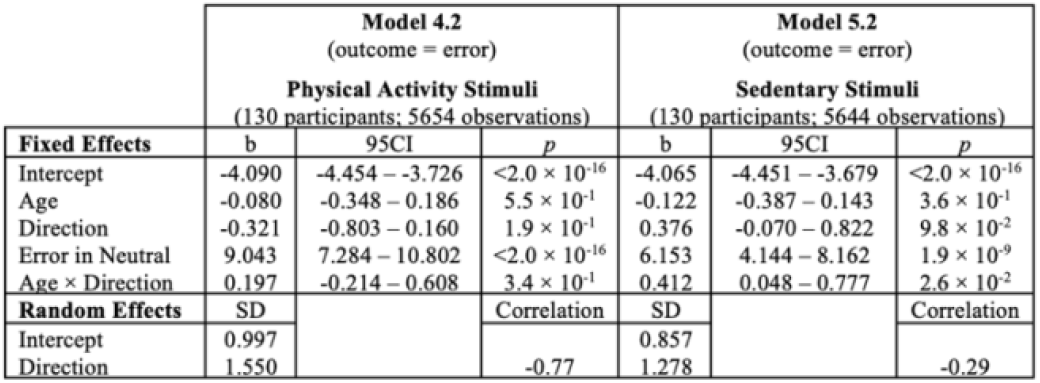
Estimated interaction effect between age and action direction on error in the physical activity (Model 4) and sedentary condition (Model 5) with mean error in the neutral condition included in the models.

**Figure 3.**
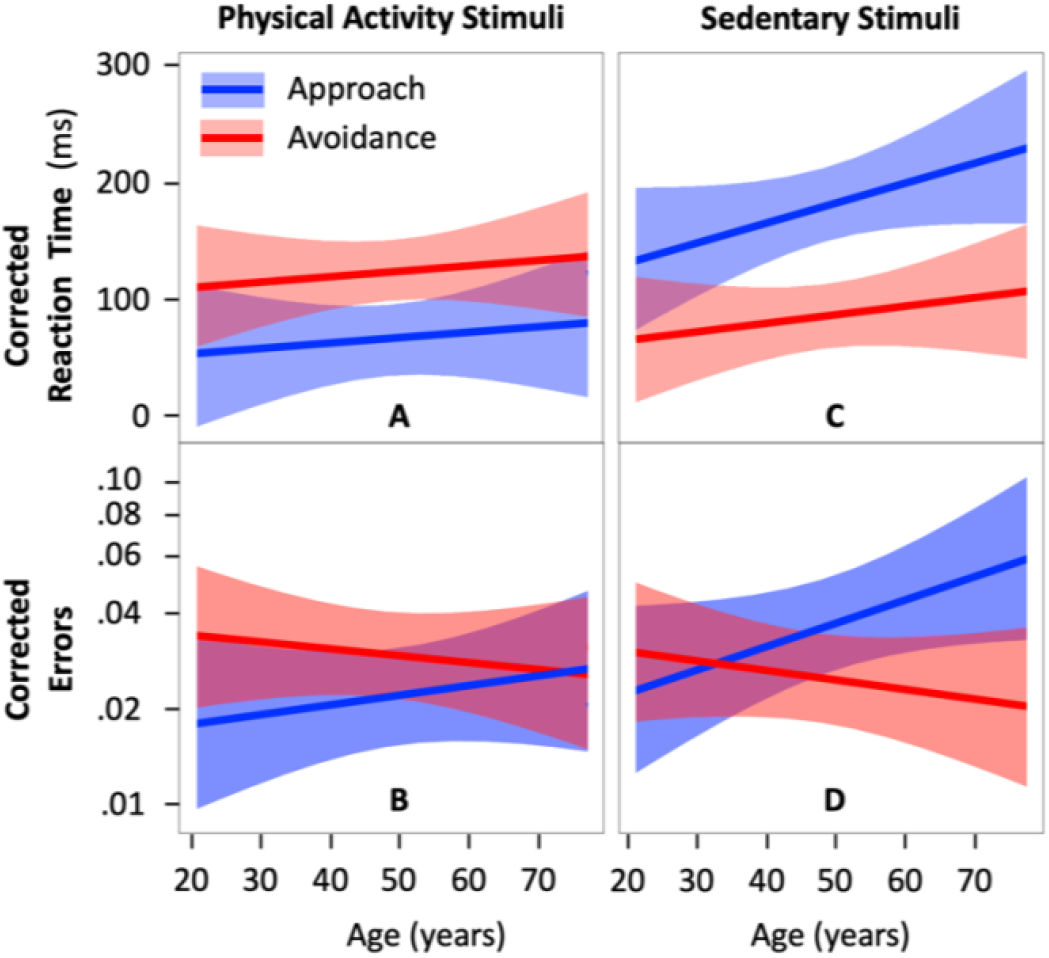
Estimation of the reaction time to approach and avoid physical activity (A) and sedentary stimuli (C) respectively corrected for the reaction time to approach and avoid neutral stimuli and the corresponding corrected errors (B and D, respectively). The colored area around the regression lines represents the 95% confidence interval.

#### Age & Sedentary Stimuli: Results Accounting for a Generic Approach-Avoidance Bias

Model 5 showed no evidence of an interaction between age and action direction on corrected reaction time to sedentary stimuli (b = 16.4; 95CI = -18.1 – 51.0; 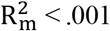 ; *p* = .353) (Table 2). However, results showed a significant main effect of action direction (b = 95.0; 95CI = 60.3 – 129.6; 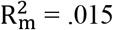 ; *p* = 3.9 × 10^−7^), with faster reactions to avoid than approach sedentary stimuli (Figure 3C).

Model 5.2 showed a significant interaction effect between age and action direction on error in the sedentary condition (b = 0.412; 95CI = 0.048 – 0.777; 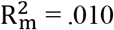 ;*p* = .026) (Table 3). A simple effect analysis suggested that the effect of age is more pronounced in the approach condition than in the avoid condition (b = 0.291; 95CI = -0.009 – 0.591; *p* = .057 vs. b = -0.122; 95CI = -0.387 – 0.143; *p* = .368) (Figure 3D). After 50 years of age, participants made significantly fewer errors when avoiding than approaching sedentary stimuli. This result is consistent with the faster reactions to avoid than approach sedentary stimuli evidenced in Model 2.

#### Sensitivity Analyses

Results of the models that did not adjust for sex, body mass index, usual physical activity, and chronic health condition were all consistent with the results reported in sections 3.2.1 to 3.2.5.

### Explicit Affective Attitude & Intention to Be Physically Active

#### Physical Activity Stimuli

Model 6 showed a significant interaction effect between explicit attitude and action direction on reaction time to physical activity stimuli (b = 95.5; 95CI = 11.3 – 179.6; 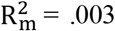 ; *p* = .028) (Figure 4A; Table 4). A simple effect analysis revealed that corrected reaction time was significantly faster in the approach condition than in the avoid condition when explicit attitude toward physical activity was the highest (pleasantness = 7; b = 127.2; 95CI = 44.3 – 209.8; *p* = .003) but showed no evidence of an effect of action direction when explicit attitude was lower (pleasantness < 7; b = 31.5; 95CI = -18.3 – 81.4; *p* = .219). Results showed no evidence of an interaction between intention and action direction on reaction time to physical activity stimuli (b = -20.2; 95CI = -103.5 – 62.9; 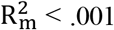 ; *p* = .635) (Figure 4B; Supplemental Table 6).

**Table 4.**
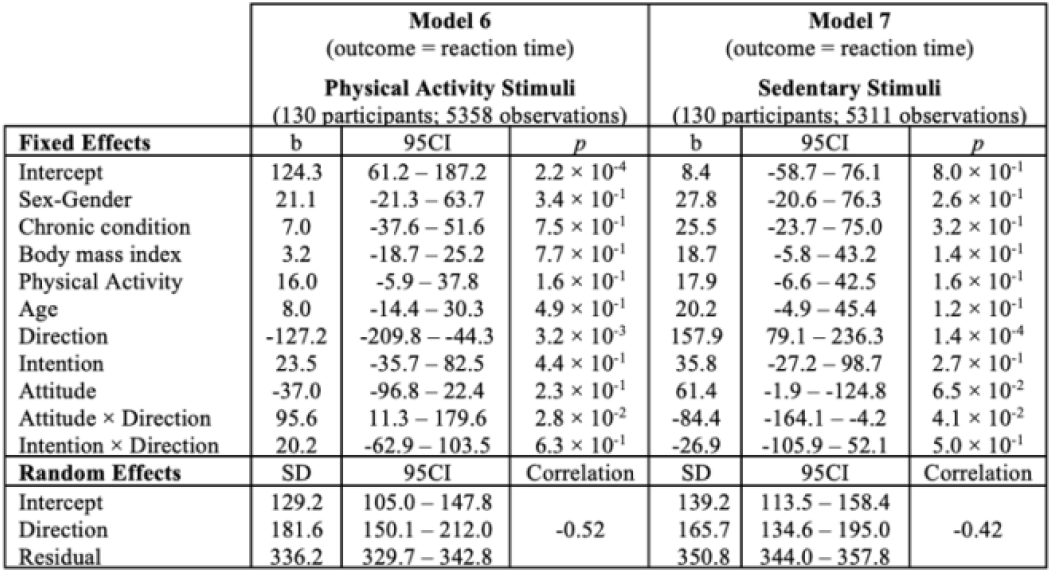
Estimated effect of explicit affective attitude and the intention to be active on corrected reaction time to approach and avoid physical activity (Model 6) and sedentary stimuli (Model 7). 95CI = 95% confidence interval, SD = standard deviation.

**Figure 4.**
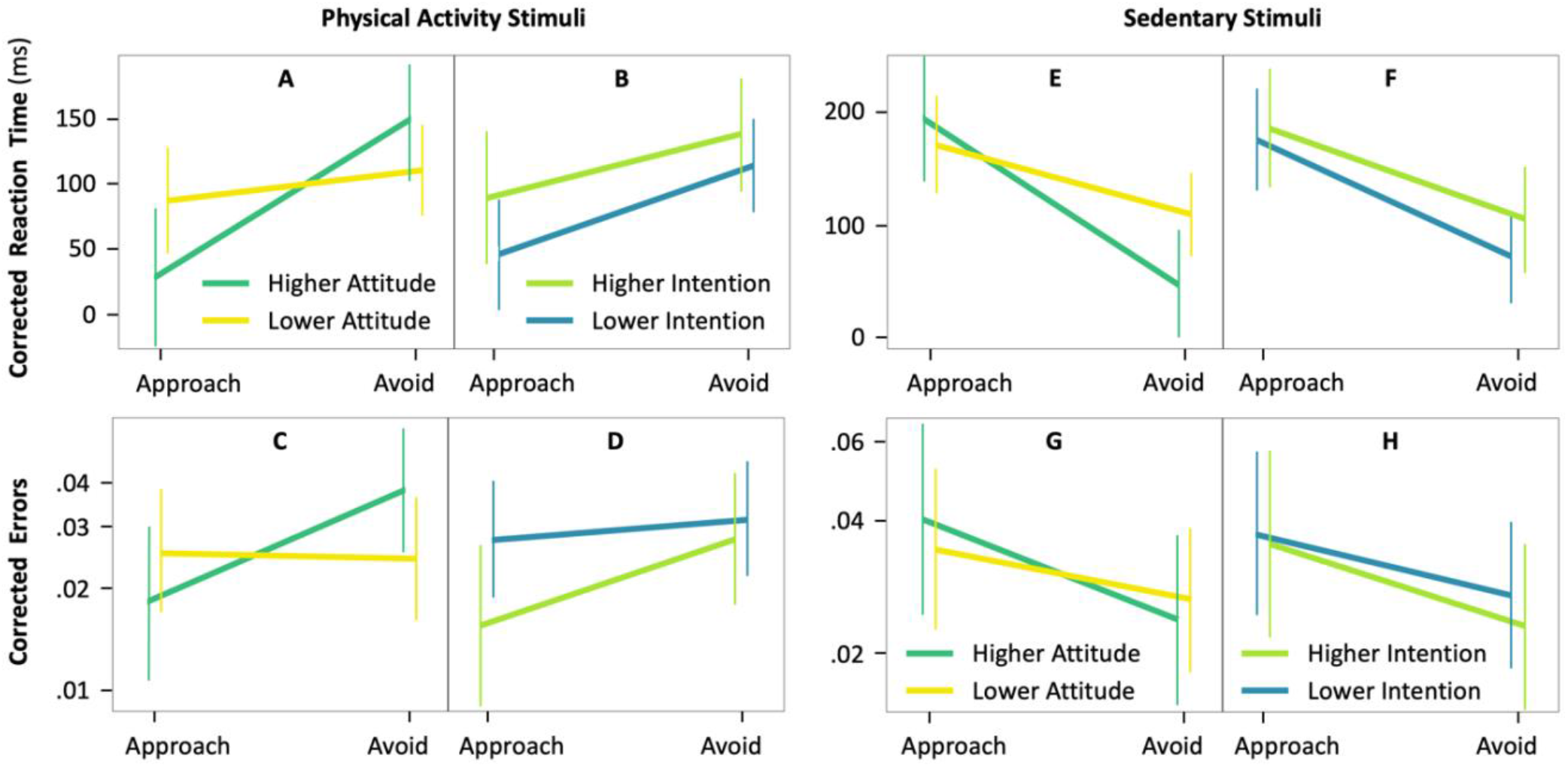
Interactions of action (approach vs. avoid) with explicit affective attitude and the intention to be physically active (right panels) on corrected reaction time (A, B, E, and F) and corrected errors (C, D, G, and H) to physical activity (left panels) and sedentary stimuli (right panels) (n = 130 participants). Higher (more positive) explicit affective attitude and higher intention correspond to a score equal 7. Lower (less positive) explicit affective attitude and lower intention correspond to a score below 7. Vertical lines are 95% confidence intervals.

Model 6.2 showed no evidence of an interaction effect between explicit attitude and action direction on error in the physical activity condition (b = -0.829; 95CI = -1.668 – 0.008; 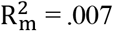 ; *p* = .052) and no evidence of an effect of explicit attitude (b = 0.358; 95CI = -0.235 – 0.952; 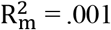 ; *p* = .237) (Figure 4C; Supplemental Table 7). Model 6.3 showed no evidence of an interaction effect between the intention to be active and action direction on error in the physical activity condition (b = 0.455; 95CI = -0.398 – 1.308; 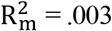 ; *p* = .295) and no evidence of an effect of intention (b = -0.589; 95CI = -1.193 – 0.013; 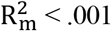 ;*p* = .055) (Figure 4D; Supplemental Table 7).

#### Sedentary Stimuli

Model 7 showed a significant interaction effect between explicit attitude and action direction on reaction time to sedentary stimuli (b = 84.4; 95CI = 4.2 – 164.1; 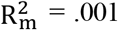 ; *p* = .041) (Figure 4E; Table 4). A simple effect analysis revealed that corrected reaction time was significantly faster in the avoid condition than in the approach condition when affective attitude toward physical activity was the highest (b = -157.9; 95CI = -236.3 – -79.1; *p* = 1.4 × 10^−4^). This significant difference was less pronounced when explicit attitude was lower (b = -73.5; 95CI = -120.7 – -26.3; *p* = .002). Results showed no evidence of an interaction effect between intention and action direction on reaction time to sedentary stimuli (b = 26.9; 95CI = -52.1 – 105.9; 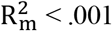 ; *p* = .506) (Figure 4F; Supplemental Table 6).

Model 7.2 showed no evidence of an interaction effect between explicit attitude and action direction on error in the sedentary condition (b = 0.271; 95CI = -0.474 – 1.017; 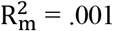 ;*p* = .475) and no evidence of an effect of explicit attitude (b = -0.107; 95CI = -0.644 – 0.429; 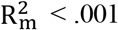 ; *p* = .695) (Figure 4G; Supplemental Table 8). Model 7.3 showed no evidence of an interaction effect between the intention to be active and action direction on error in the sedentary condition (b = 0.118; 95CI = -0.633 – 0.869; 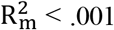; *p* = .758) and no evidence of an effect of intention (b = -0.176; 95CI = -0.709 – 0.356; 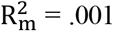 ; *p* = .517) (Figure 4H; Supplemental Table 8).

#### Sensitivity Analyses

When Model 6 and Model 7 were not adjusted for sex-gender, body mass index, usual physical activity, and chronic health condition, the results remained consistent with the main results reported above. Moreover, the significance of the interactions involving explicit attitude or intention remained consistent with the main results when these interactions were tested in separate models.

## Discussion

### Main Findings

Our results confirmed a main tendency to approach physical activity stimuli (i.e., faster reaction to approach vs. avoid) and to avoid sedentary stimuli (i.e., faster reaction to avoid vs. approach) across the lifespan. Importantly, results based on neutral stimuli revealed a generic approach tendency in early adulthood (i.e., faster approach before age 53 and fewer errors before age 36) and a generic avoidance tendency in older adults (i.e., more errors after age 60). Contrary to our preregistered hypotheses, when accounting for these generic tendencies, our results showed a greater tendency (i.e., fewer errors) to avoid than approach sedentary stimuli after age 50, but not before, and no evidence of an effect of age on approach-avoidance tendencies toward physical activity stimuli. Finally, exploratory analyses showed that, irrespective of age, participants were faster at approaching physical activity and avoiding sedentary stimuli when they considered physical activity as pleasant and enjoyable (explicit attitude). However, results showed no evidence of an association between approach-avoidance tendencies and the intention to be physically active. Taken together, these results suggest that both age and explicit attitudes can affect the general tendency to approach physical activity stimuli and to avoid sedentary stimuli.

### Comparison With the Literature

Our results showing an age-related decline in generic approach bias are consistent with an 8-year longitudinal personality study showing that self-reported approach motivation was the highest in younger adults and the lowest in older adults (Windsor et al., 2012). Further, an average intra-individual decline was evidenced in an 8-year period in younger, middle-aged, and older adults (Windsor et al., 2012). Taken together, these consistent findings may reflect the changing orientation of personal goals over the course of adulthood, beginning with the pursuit of growth in young adults, to maintenance in adults, and the prevention of loss in older adults (Ebner et al., 2006). Because our results suggest a generic approach-avoidance bias in younger and older adults, future studies testing approach-avoidance tendencies in these populations should control for this potential confounder.

Both reaction-time and error results supported a tendency to approach physical activity stimuli and to avoid sedentary stimuli. These results are consistent with previous literature (Cheval et al., 2014; Cheval et al., 2015; Cheval et al., 2016a; Cheval et al., 2018c; Hannan et al., 2019; Moffit et al., 2019). Our study extends these previous results by showing that these biases apply to all adult ages and that the avoidance bias for sedentary stimuli increases with aging. According to the effortless self-control hypothesis arguing that individuals are faster to approach (vs. avoid) their long-term goals and to avoid (vs. approach) temptations (Fishbach & Shah, 2006), these results suggest that the guidelines published for over two decades (World Health Organization, 1996; World Health Organization, 2020) aiming to promote physical activity have been successful in developing physical activity as a long-term goal. They also suggest that sedentary behavior is a temptation, which is consistent with recent theoretical and experimental work on the rewarding value of effort minimization (Cheval et al., 2018a) and its automatic attraction (Boisgontier & Iversen, 2020; Cheval et al., 2020a; Cheval & Boisgontier, 2021; Cheval et al., 2021a).

Our error-based results showing an association between automatic approach-avoidance tendencies toward sedentary stimuli and explicit affective attitudes toward physical activity are consistent with previous results based on reactions times showing an association of automatic attitudes toward physical activity with explicit affective attitudes (Muschalik et al., 2019). Moreover, our results do not contradict previous results based on reaction times showing no evidence of such associations (Hyde et al., 2010; Rebar et al., 2015; Sala et al., 2016; Muschalik et al., 2018) since these previous studies did not investigate errors as an outcome. Taken together, these results further support recent findings suggesting that automatic and explicit attitudes are not independent (Greene et al., 2001; Hofmann et al., 2005; Nosek, 2005; Béna et al., 2022). Future studies are needed to examine moderators of the association between these implicit and explicit constructs in the physical activity domain (e.g., Berry et al., 2016).

The absence of evidence supporting an association between automatic attitudes and intentions to be physically active is consistent with previous studies (Conroy et al., 2010; Cheval et al., 2014; Rebar et al., 2015; Muschalik et al., 2019; Hannan et al., 2021). However, our results are inconsistent with the study by Banting et al. (2009) and Cheval et al. (2015). This discrepancy may be explained by the fact that previous studies did not account for a potential generic approach-avoidance tendency that could vary across participants, potentially confounding the results. Further, the possibility that these effects on reaction times are in fact counterbalanced by inverse effects on errors cannot be discounted since these errors were not analyzed in these studies.

Considering the intertwining of emotions, approach-avoidance tendencies, and behavior (Lang, 1995), future studies should go beyond the measure of motivational direction (approach vs. avoid) generated by the stimuli. Coupling the approach-avoidance task with self-reported (e.g., self-assessment manikin) (Bradley and Lang, 1994) and other behavioral measures (e.g., eye-tracking) to investigate other indicators (i.e., stimulus-generated arousal and valence) could allow for a better understanding of the automatic reactions associated with physical activity and sedentary stimuli and how they relate to behavioral regulation in a specific context (Moors et al., 2013).

### Limitations & Strengths

The present study has potential limitations. First, the online nature of the study made it impossible to limit the influence of potential distractions in the participant’s environment and to control whether participants were using their two index fingers to perform the task as instructed and whether they were sitting or standing, which could have influenced the results (Cheval et al., 2018a). Second, the data were mainly collected in Canada and France. It is thus unclear whether conclusions could generalize to populations from non-Western countries or less active populations of older adults. Third, the older adults of our sample were more active than the young adults, which may result from recruitment bias. Although we controlled for this potential bias by including the usual level of physical activity in the models testing the effect of age, whether conclusions generalize to a sample with less active older adults would need to be confirmed. Fourth, the usual level of physical activity was assessed using a self-reported questionnaire, which may not accurately reflect the objective level of physical activity. Assessing physical activity and sedentary behaviors using device-based measures would have provided a more reliable estimate.

Moreover, the current study did not assess the potential influence of socioeconomic (e.g., income, education; see Pechey et al., 2015 in the food domain), the quality of the motivation towards physical activity (i.e., autonomous vs. controlled) (Berry et al., 2016), personality, and goal-related variables (Elliot, 1999; Elliot et al., 2002) on automatic approach-avoidance tendencies. Testing these associations in future work would clarify the mechanisms underlying the effect of age on approach-avoidance tendencies (e.g., moderating effect of income on the association between age and approach-avoidance tendencies). Regarding motivation quality, beyond developing the intention to be physically active, it seems important to disentangle the reasons beyond this intention (e.g., from more intrinsic to more extrinsic reasons) and to examine how individuals’ reasons for action may correlate with more automatic constructs.

However, these limitations are counterbalanced by several strengths. Among these strengths are a preregistered hypothesis (Boisgontier, 2021) and a sample size based on an a priori power analysis, which are considered good research practices (Caldwell et al., 2020; Boisgontier, 2022). In addition, as recommended in a critical review of measurement practices in the study of automatic associations of physical activity and sedentary behavior (Zenko & Ekkekakis, 2019a), we justify the choice of the approach-avoidance task and report moderate to good internal consistency of both reaction-time and error bias for each type of stimuli. Other strong points include an objective measure of automatic attitudes, accounting for a generic approach-avoidance bias that could have confounded the results, the use of statistics limiting information loss (i.e., mixed-effects models), and consistent results across the two outcomes (i.e., reaction time and errors) as well as across main and sensitivity analyses.

## Data Availability

All data produced are available online at https://doi.org/10.5281/zenodo.7050947

https://doi.org/10.5281/zenodo.7050947

## Additional Information

### Data & Code Availability

All supplemental material, code, and data are freely available on the Zenodo open-access repository: https://doi.org/10.5281/zenodo.7050947 (Farajzadeh et al., 2022).

### CRediT Authorship Contribution Statement

(Brand et al., 2015; Allen et al., 2019)

- **Ata Farajzadeh**: Writing – Review & Editing.
- **Miriam Goubran**: Writing – Review & Editing.
- **Alexa Beehler**: Investigation.
- **Noura Cherkaoui**: Investigation.
- **Paula Morrison**: Investigation.
- **Margaux de Chanaleilles**: Software.
- **Silvio Maltagliati**: Writing – Review & Editing.
- **Boris Cheval**: Writing – Review & Editing.
- **Matthew W. Miller**: Writing – Review & Editing.
- **Lisa Sheehy**: Writing – Review & Editing.
- **Martin Bilodeau**: Writing – Review & Editing.
- **Dan Orsholits**: Formal Analysis (Supporting), Data Curation.
- **Matthieu P. Boisgontier**: Conceptualization, Methodology, Software, Formal Analysis (Lead), Investigation, Resources, Data Curation, Writing – Original Draft, Visualization, Supervision, Project Administration, Funding Acquisition.

