## Supplemental Material for "Automatic approach-avoidance tendency toward physical activity, sedentary, and neutral stimuli as a function of age, explicit affective attitude, and intention to be active" (from another version of this preprint: version 1, 2022-09-09)

MedRxiv, 2022

**Supplemental Figure 1.** Intention to be physically active.

**Supplemental Figure 2.** Explicit affective attitude toward physical activity.

**Supplemental Figure 3.** Distribution of the participants per 10-year age ranges.

**Supplemental Table 1.** Observations, reaction time > 3,000 ms, reaction time < 150 ms, and error as a function of stimuli and action direction.

**Supplemental Table 2**. Correlation table.

**Supplemental Table 3.** Estimated interaction effect between age and action direction on the reaction time to physical activity (Model 1) and sedentary stimuli (Model 2).

**Supplemental Table 4.** Estimated interaction effect between age and action direction on error in the physical activity (Model 1.2) and sedentary condition (Model 2.2).

**Supplemental Table 5.** Estimated interaction effect between age and action direction on the reaction time to neutral stimuli (Model 3).

**Supplemental Table 6.** Estimated interaction effect between age and action direction on error in the neutral condition (Model 3.2).

**Supplemental Table 7.** Estimated effect of explicit affective attitude (Model 6.2) and intention to be active (Model 7.2) on error when reacting to physical activity stimuli.

**Supplemental Table 8.** Estimated effect of explicit affective attitude (Model 6.3) and intention to be active (Model 7.3) on error when reacting to sedentary stimuli.

**Supplemental Figure 1.** Intention to be physically active.

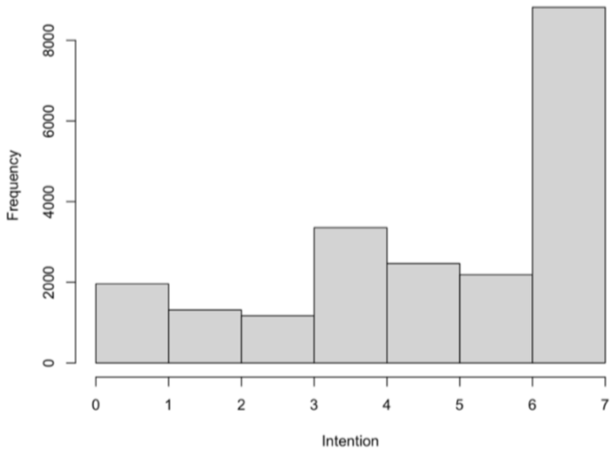


**Supplemental Figure 2.** Explicit affective attitude toward physical activity.

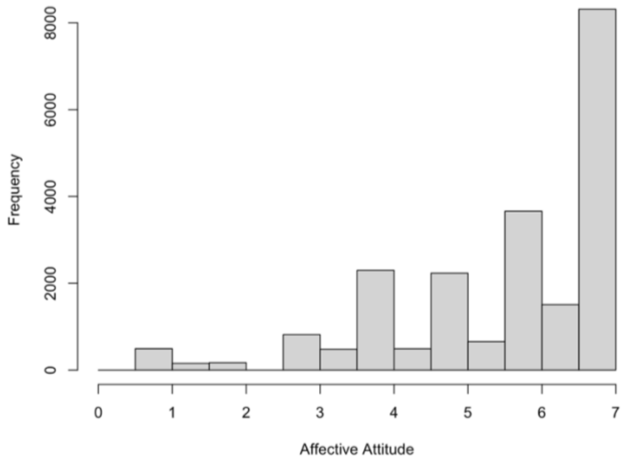


**Supplemental Figure 3.** Distribution of the participants per 10-year age ranges.

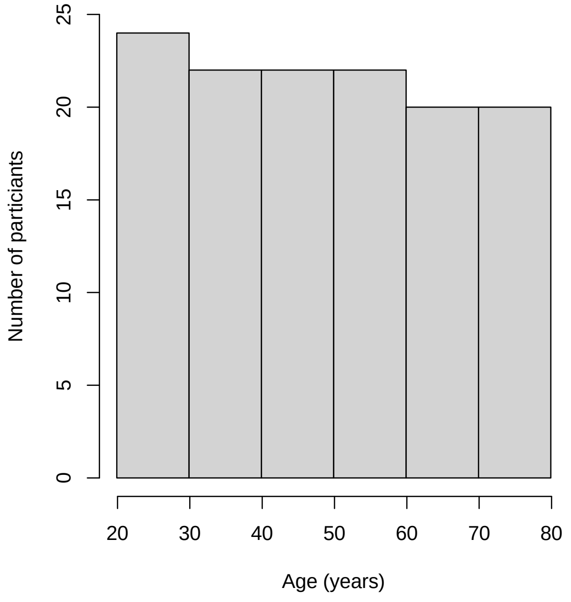

**Supplemental Table 1.** Observations, reaction time > 3,000 ms, reaction time < 150 ms, and error as a function of stimuli and action direction.

| Stimuli | Action | Total number of observations | Reaction time > 3,000 ms | Reaction time < 150 ms | Errors |
|---|---|---|---|---|---|
| Physical activity | Approach | 2917 | 96 (3.2 %) | 1 (<0.1 %) | 162 (5.5 %) |
| | Avoid | 2944 | 109 (3.7 %) | 1 (<0.1 %) | 179 (6.0 %) |
| Sedentary | Approach | 2940 | 149 (5.0 %) | 1 (<0.1 %) | 245 (8.3 %) |
| | Avoid | 2953 | 97 (3.2 %) | 2 (<0.1 %) | 137 (4.3 %) |
| Neutral | Approach | 5196 | 161 (3.0 %) | 31 (0.5 %) | 400 (7.6 %) |
| | Avoid | 5139 | 153 (2.9 %) | 22 (0.4 %) | 361 (7.0 %) |
| **Total** | | **22,089** | **765 (3.4 %)** | **58 (0.2 %)** | **1484 (6.7 %)** |

**Supplemental Table 2.** Correlation table. *p < .05

| | 1 | 2 | 3 | 4 | 5 | 6 | 7 | 8 | 9 | 10 | 11 |
|---|---|---|---|---|---|---|---|---|---|---|---|
| 1. Physical activity stimuli – Approach | – | 0.44 | 0.51 | 0.52 | 0.45 | 0.46 | 0.48 | -0.08 | -0.08 | 0.12 | 0.01 |
| 2. Physical activity stimuli – Avoid | 0.44 | – | 0.52 | 0.44 | 0.44 | 0.42 | 0.41 | 0.00 | -0.02 | 0.08 | -0.02 |
| 3. Sedentary stimuli – Approach | 0.51 | 0.52 | – | 0.49 | 0.47 | 0.45 | 0.46 | -0.03 | -0.07 | 0.10 | -0.01 |
| 4. Sedentary stimuli – Avoid | 0.52 | 0.44 | 0.49 | – | 0.47 | 0.45 | 0.44 | -0.07 | -0.07 | 0.09 | 0.04 |
| 5. Neutral stimuli – Approach | 0.45 | 0.44 | 0.47 | 0.47 | – | 0.49 | 0.45 | -0.04 | -0.08 | 0.07 | -0.02 |
| 6. Neutral stimuli – Avoid | 0.46 | 0.42 | 0.45 | 0.45 | 0.49 | – | 0.40 | -0.02 | -0.07 | 0.06 | -0.00 |
| 7. Age | 0.48 | 0.41 | 0.46 | 0.44 | 0.45 | 0.40 | – | -0.10 | -0.13 | 0.22 | 0.06 |
| 8. Explicit affective attitude | -0.08 | 0.00 | -0.03 | -0.07 | -0.04 | -0.02 | -0.10 | – | 0.61 | 0.16 | -0.25 |
| 9. Intention to be active | -0.08 | -0.02 | -0.07 | -0.07 | -0.08 | -0.07 | -0.13 | 0.61 | – | 0.24 | -0.20 |
| 10. Usual level of physical activity | 0.12 | 0.08 | 0.10 | 0.09 | 0.07 | 0.06 | 0.22 | 0.16 | 0.24 | – | -0.14 |
| 11. Body mass index | 0.01 | -0.02 | -0.01 | 0.04 | 0.02 | -0.00 | 0.06 | -0.25 | -0.20 | -0.14 | – |

**Supplemental Table 3.** Estimated interaction effect of age and action direction on the reaction time to physical activity (Model 1) and sedentary stimuli (Model 2). 95CI = 95% confidence interval, SD = standard deviation.

| | **Model 1** (outcome = reaction time) **Physical Activity Stimuli** (130 participants; 5358 observations) | | | **Model 2** (outcome = reaction time) **Sedentary Stimuli** (130 participants; 5311 observations) | | |
|---|---|---|---|---|---|---|
| **Fixed Effects** | b | 95CI | *p* | b | 95CI | *p* |
| Intercept | 1170.3 | 1097.4 – 1243.0 | $2.0 \times 10^{-16}$ | 1108.6 | 1036.0 – 1181.3 | $2.0 \times 10^{-16}$ |
| Sex-Gender | -175.0 | -264.5 – -85.4 | $2.3 \times 10^{-4}$ | -140.9 | -233.5 – -48.4 | $2.1 \times 10^{-3}$ |
| Chronic condition | -57.9 | -151.7 – 35.7 | $2.2 \times 10^{-1}$ | -42.8 | -135.8 – 50.2 | $4.5 \times 10^{-1}$ |
| Body mass index | -6.7 | -51.6 – 38.2 | $7.7 \times 10^{-1}$ | 16.3 | -28.4 – 61.3 | $9.3 \times 10^{-1}$ |
| Physical Activity | 6.8 | -38.2 – 51.9 | $7.6 \times 10^{-1}$ | 3.2 | -42.1 – 48.4 | $8.8 \times 10^{-1}$ |
| Age | 190.4 | 141.2 – 239.5 | $1.1 \times 10^{-11}$ | 195.1 | 147.5 – 242.7 | $9.6 \times 10^{-13}$ |
| Direction | -88.0 | -121.2 – -54.6 | $8.9 \times 10^{-7}$ | 68.8 | 33.3 –104.5 | $1.4 \times 10^{-4}$ |
| Age × Direction | 39.9 | 6.7 – 73.1 | $2.0 \times 10^{-2}$ | 56.9 | 21.4 – 92.6 | $2.0 \times 10^{-3}$ |
| **Random Effects** | SD | 95CI | Correlation | SD | 95CI | Correlation |
| Intercept | 260.3 | 223.1 – 291.6 | | 248.8 | 212.6 – 278.6 | |
| Direction | 161.4 | 132.4 – 190.8 | -0.26 | 174.4 | 143.6 – 205.6 | -0.01 |
| Residual | 336.2 | 329.7 – 342.8 | | 350.6 | 343.9 – 357.5 | |

**Supplemental Table 4.** Estimated interaction effect between age and action direction on error in the physical activity (Model 1.2) and sedentary condition (Model 2.2).

| | **Model 1.2** (outcome = error) **Physical Activity Stimuli** (130 participants; 5654 observations) | | | **Model 2.2** (outcome = error) **Sedentary Stimuli** (130 participants; 5644 observations) | | |
|---|---|---|---|---|---|---|
| **Fixed Effects** | b | 95CI | *p* | b | 95CI | *p* |
| Intercept | -3.541 | -3.914 – -3.169 | $<2.0 \times 10^{-16}$ | -3.697 | -4.068 – -3.326 | $<2.0 \times 10^{-16}$ |
| Age | -0.007 | -0.318 – 0.304 | $9.6 \times 10^{-1}$ | 0.115 | -0.177 – 0.409 | $4.4 \times 10^{-1}$ |
| Direction | -0.383 | -0.908 – 0.141 | $1.5 \times 10^{-1}$ | 0.422 | -0.025 – 0.870 | $6.4 \times 10^{-2}$ |
| Age × Direction | 0.500 | 0.090 – 0.911 | $1.6 \times 10^{-2}$ | 0.201 | -0.127 – 0.530 | $2.2 \times 10^{-1}$ |
| **Random Effects** | SD | | Correlation | SD | | Correlation |
| Intercept | 1.330 | | | 1.193 | | |
| Direction | 1.473 | | -0.46 | 1.050 | | -0.20 |

**Supplemental Table 5.** Estimated interaction effect of age and action direction on the reaction time to neutral stimuli (Model 3). 95CI = 95% confidence interval, SD = standard deviation.

| | **Model 3** (outcome = reaction time) **Neutral Stimuli** (130 participants; 9302 observations) | | |
|---|---|---|---|
| **Fixed Effects** | b | 95CI | *p* |
| Intercept | 1061.1 | 995.1 – 1127.3 | $2.0 \times 10^{-16}$ |
| Sex-Gender | -207.9 | -290.4 – -125.3 | $2.4 \times 10^{-6}$ |
| Chronic condition | -66.2 | -150.6 – 18.1 | $1.3 \times 10^{-1}$ |
| Body mass index | -11.3 | -52.2 – 29.5 | $5.8 \times 10^{-1}$ |
| Physical Activity | -13.3 | -54.1 – 27.4 | $5.2 \times 10^{-1}$ |
| Age | 175.0 | 131.1 – 218.9 | $3.7 \times 10^{-12}$ |
| Direction | -31.0 | -48.8 – -13.4 | $7.8 \times 10^{-4}$ |
| Age × Direction | 40.7 | 23.1 – 58.3 | $1.5 \times 10^{-5}$ |
| **Random Effects** | SD | 95CI | Correlation |
| Intercept | 233.9 | 201.0 – 261.3 | |
| Direction | 60.0 | 33.4 – 81.3 | -0.18 |
| Residual | 350.2 | 345.1 – 355.3 | |

**Supplemental Table 6.** Estimated interaction effect between age and action direction on error in the neutral condition (Model 3.2).

| | **Model 3.2** (outcome = error) **Physical Activity Stimuli** (130 participants; 9968 observations) | | |
|---|---|---|---|
| **Fixed Effects** | b | 95CI | *p* |
| Intercept | -3.079 | -3.311 – -2.847 | $<2.0 \times 10^{-16}$ |
| Age | -0.032 | -0.241 – 0.183 | $7.6 \times 10^{-1}$ |
| Direction | 0.021 | -0.209 – 0.251 | $8.5 \times 10^{-1}$ |
| Age × Direction | 0.391 | 0.200 – 0.582 | $6.0 \times 10^{-5}$ |
| **Random Effects** | SD | | Correlation |
| Intercept | 1.010 | | |
| Direction | 1.485 | | -0.19 |

**Supplemental Table 7.** Estimated effect of explicit affective attitude (Model 6.2) and intention to be active (Model 7.2) on error when reacting to physical activity stimuli.

| | **Model 6.2** (outcome = error) **Physical Activity Stimuli** (130 participants; 5654 observations) | | | **Model 6.3** (outcome = error) **Physical Activity Stimuli** (130 participants; 5654 observations) | | |
|---|---|---|---|---|---|---|
| **Fixed Effects** | b | 95CI | *p* | b | 95CI | *p* |
| Intercept | -4.617 | -5.183 – -4.051 | $<2.0 \times 10^{-16}$ | -4.171 | -4.613 – -3.729 | $<2.0 \times 10^{-16}$ |
| Attitude | 0.358 | -0.235 – 0.952 | $2.3 \times 10^{-1}$ | | | |
| Intention | | | | -0.589 | -1.193 – 0.013 | $5.5 \times 10^{-2}$ |
| Direction | 0.769 | 0.095 – 1.442 | $2.5 \times 10^{-2}$ | 0.134 | -0.433 – 0.703 | $6.4 \times 10^{-1}$ |
| Error in Neutral | 9.186 | 7.487 – 10.884 | $<2.0 \times 10^{-16}$ | 9.059 | 7.409 – 10.708 | $<2.0 \times 10^{-16}$ |
| Attitude × Direction | -0.829 | -1.668 – 0.008 | $5.2 \times 10^{-2}$ | | | |
| Intention × Direction | | | | 0.455 | -0.398 – 1.308 | $2.9 \times 10^{-1}$ |
| **Random Effects** | SD | | Correlation | SD | | Correlation |
| Intercept | 1.009 | | | 0.993 | | |
| Direction | 1.546 | | -0.78 | 1.557 | | -0.79 |

**Supplemental Table 8.** Estimated effect of explicit affective attitude (Model 6.3) and intention to be active (Model 7.3) on error when reacting to sedentary stimuli.

| | **Model 6.3** (outcome = error) **Sedentary Stimuli** (130 participants; 5654 observations) | | | **Model 7.3** (outcome = error) **Sedentary Stimuli** (130 participants; 5654 observations) | | |
|---|---|---|---|---|---|---|
| **Fixed Effects** | b | 95CI | *p* | b | 95CI | *p* |
| Intercept | -4.005 | -4.434 – -3.576 | $<2.0 \times 10^{-16}$ | -3.970 | -4.420 – -3.520 | $<2.0 \times 10^{-16}$ |
| Attitude | -0.107 | -0.644 – 0.429 | $6.9 \times 10^{-1}$ | | | |
| Intention | | | | -0.176 | -0.709 – 0.356 | $5.1 \times 10^{-1}$ |
| Direction | 0.271 | -0.277 – 0.819 | $3.3 \times 10^{-1}$ | 0.331 | -0.217 – 0.880 | $2.3 \times 10^{-1}$ |
| Error in Neutral | 5.925 | 3.936 – 7.914 | $5.2 \times 10^{-9}$ | 5.852 | 3.890 – 7.813 | $5.0 \times 10^{-9}$ |
| Attitude × Direction | 0.271 | -0.474 – 1.017 | $4.7 \times 10^{-1}$ | | | |
| Intention × Direction | | | | 0.118 | -0.633 – 0.869 | $7.5 \times 10^{-1}$ |
| **Random Effects** | SD | | Correlation | SD | | Correlation |
| Intercept | 0.853 | | | 0.849 | | |
| Direction | 1.314 | | -0.29 | 1.320 | | -0.29 |
